# Evaluating tuberculosis treatment outcomes and predictors in five Southern African countries: A multi-country cohort analysis

**DOI:** 10.64898/2026.03.18.26348675

**Authors:** Mirriam Ndhlovu, Leticia Wuethrich, Jacqueline Huwa, Agness Thawani, Geldert Chiwaya, Aubrey Kudzala, Joseph Chintedza, Guy Muula, Denise Evans, Idiovino Rafael, Cordelia Kunzekwenyika, Fiona Mureithi, Nelly Jinga, Amina Fernando, Marie Ballif, Gunar Günther, Lukas Fenner, Nicolas Banholzer, IeDEA Southern Africa (IeDEA-SA)

**Author notes:** Corresponding authors: Mirriam Ndhlovu, Prof. Lukas Fenner. These authors contributed equally to this work (Mirriam Ndhlovu, Leticia Wuethrich). These authors also contributed equally to this work (Lukas Fenner, Nicolas Banholzer).

## Abstract

**Introduction:** Despite global progress in tuberculosis (TB) control, treatment outcomes remain suboptimal, particularly in high-burden settings and among people with HIV or drug-resistant TB. Identifying predictors of unsuccessful treatment is essential to improve TB care and policy.

**Methods:** We evaluated TB treatment outcomes and patient characteristics associated with unsuccessful outcomes in five cohorts of the International epidemiology database to evaluate AIDS (IeDEA); Center for Infectious Disease Research, Zambia; Chiure health center, Mozambique; Martin Preuss Center, Lighthouse clinic, Malawi; Masvingo health center Zimbabwe; and Themba Lethu clinic, Hellen Joseph hospital, South Africa. We included all patients with TB aged ≥ 15 years starting TB treatment and assessed their treatment outcomes in association with sociodemographic and clinical characteristics using multivariable mixed-effects models. Unsuccessful outcomes were defined as death, loss to follow-up and treatment failure.

**Results:** Among 1438 people with TB, median age was 39 years, 67% males, 40% with HIV, and 4% with MDR-TB; 1151 (80%) treatment outcomes were successful (606 cured and 545 completed treatment), 221 (15%) unsuccessful (89 deaths, 129 loss to follow-up and 3 treatment failures), and 66 (5%) other (49 unknown and 17 transfer-outs). Unsuccessful outcomes were more probable among people with multidrug-resistant TB (MDR-TB) and among participants without formal education. Risk of death was lower for people with bacteriologically confirmed TB (adjusted odds ratio (aOR) 0.5, 95%-credible interval [CI] 0.25-0.80), those with a secondary or higher education (aOR 0.3, 95%-CI 0.13-0.69) and BMI ≥18 kg/m² (aOR 0.6, 95%-CI 0.36-0.99). MDR-TB was associated with an increase (aOR 2.4 95%-CI 1.17-4.97) and primary and secondary or higher education with a decrease in loss to follow-up (aOR 0.3, 95%-CI 0.14-0.89 and aOR 0.3, 95%-CI 0.11-0.67, respectively).

**Conclusions:** TB treatment outcomes fell short of the targets set by the World Health Organization of <10% unsuccessful outcomes, indicating a critical need for enhanced management strategies. Tackling loss to follow-up is crucial, especially among MDR-TB patients, including stronger retention activities and improved diagnostic capacities.

## Introduction

Tuberculosis (TB) is the leading cause of death among infectious diseases (1). Low and middle income countries experience a high burden of TB especially in the Southern African region (2). The strategies to end TB include the reduction of the number of deaths by 95%, and increasing cure rates among patients receiving first line treatment to 90% between 2015 and 2035 (3). Although the global TB treatment success rate among individuals treated with first line TB regimens increased from 80.1% in 2019 to 88% in 2024, the success rate among people with Multi Drug Resistant/Rifampicin Resistant TB (MDR/RR-TB) was still only 71% (4). Furthermore, treatment success rate for Extensively Drug Resistant (XDR) TB was 34% in 2018 (5).

The World Health Organization (WHO) African region had 48% reduction in TB related deaths between 2015 to 2025 (4). Although significant advancements have been made for TB management in Southern Africa following the rollout of integrated patient centered care (3), TB treatment success rates remain below the 90% standards set by WHO. In sub-Saharan Africa, the majority of unsuccessful outcomes were deaths (48%) and loss to follow-up (47%) (6) HIV infection and TB history were consistently associated with such outcomes, but also other predictors have been reported, including age, race, low education level, illicit drug use, and alcohol consumption (7). This study evaluated TB treatment outcomes and determined factors associated with poor outcomes among people treated for TB in the Southern African region between 2022 to 2025.

## Methods

### Study design setting and timeframe

In a prospective, non-interventional cohort study, we consecutively enrolled people with pulmonary TB from five HIV care clinics and their associated TB clinics in the International epidemiology Databases to Evaluate AIDS in Southern Africa (IeDEA-SA): Center for infectious disease research in Zambia, Lusaka (Zambia), Themba Lethu Clinic, Johannesburg (South Africa), Martin Preuss Center, Lilongwe (Malawi), Chiure health Center, Chiure (Mozambique), and Masvingo health Center, Masvingo (Zimbabwe). The study sites were hospitals at primary and secondary care levels in both urban and rural areas (8).

Between October 2022 and March 2023, we recruited patients diagnosed with pulmonary TB aged ≥15 years, with or without HIV, and with or without additional extrapulmonary manifestations (e.g. lymph nodes, pleura). We included the cohort data available on March 31, 2025. Pulmonary TB was either bacteriologically confirmed through MTB GeneXpert (MTB RIF or ULTRA), microscopy, chest Xray or based on clinical presentation. All participants started TB treatment (baseline) and were followed up until the end of TB treatment. TB and associated comorbidities such as HIV were managed according to country-specific national guidelines and those set by the WHO (8). All people with TB and HIV were put on treatment. TB treatment outcomes and risk factors were collected through standardized questionnaires and entered electronically into a REDCap database (9) based on all data available and collected during the study visits (patient charts, TB registers, study investigations).

### Data collection and definitions

#### TB treatment outcomes

Treatment outcomes were defined according to WHO guidelines as successful, unsuccessful and other outcomes (unknown or transferred out) (10,11). Successful outcomes included participants who were cured or completed TB treatment. Unsuccessful outcomes included death, loss to follow-up, and treatment failures. Participants who were loss to follow-up were categorized internally if they did not return for the scheduled end-of treatment visit within six months for drug-susceptible TB or twelve months for MDR-TB, allowing a waiting period of eight months (or sixteen months for MDR-TB). Treatment failure was defined as a clinically relevant discontinuation or change of regimen due to poor clinical or bacteriological response. Other outcomes included patients who were transferred out of participating clinics or had unknown outcomes. We also recorded any non-fatal serious adverse events during treatment.

#### Predictors of TB treatment outcomes

We collected sociodemographic and clinical characteristics at baseline, including age, sex, body mass index (BMI), education level, location of residence, TB history, HIV infection, MDR-TB, and type of TB diagnosis. BMI was categorized as underweight (<18 kg/m²) or normal (18–30 kg/m²), as only few participants had BMI>30. Education level was categorized as no formal education, primary education, and secondary or higher education. Location of residence was dichotomized as rural or non-rural by combining peri-urban and urban. Substance use was assessed with the Alcohol, Smoking and Substance Involvement Screening Test (ASSIST) and categorized according to the WHO guidelines as either low risk (alcohol: 0–10; smoking/other substances: 0–3) or moderate risk (alcohol: 11–26; smoking/other substances: 4–26), as high-risk use (>26) was not observed. Since other substances were rare, we created a composite variable referring to a low or moderate risk for any use of cannabis, cocaine, inhalants, sedatives, hallucinogens, opioids, or other drugs. MDR-TB was defined as resistance to at least rifampicin. Bacteriologically confirmed TB was defined by a positive urine LAM culture, Xpert MTB/RIF assay or microscopy smear, whereas clinically diagnosed TB was based on clinical or radiographic findings without laboratory confirmation.

### Statistical Analyses

We first described TB treatment outcomes and baseline patient characteristics, which were summarized as counts and percentages. Associations were estimated using multivariable logistic regression models with country fixed effects. Patients with unknown outcomes were excluded, and those transferred out were considered alive and not loss to follow-up. Missing patient characteristics were handled using multiple imputation by chained equations (10 imputations) based on TB treatment outcomes and baseline patient characteristics. Estimates were pooled using Rubin’s rules and presented as adjusted odds ratios (aOR) with 95% confidence intervals (CIs). To describe the full distribution of treatment outcomes, we additionally fitted a multinomial logistic regression model with country fixed effects. We calculated predictive margins for each outcome by setting a specific covariate level for the entire sample while maintaining observed values for all other variables, then averaging the resulting probabilities. All analyses were performed in R (version 4.5.1). Multinomial models were fitted using the nnet (version 7.3-20) packages.

### Ethics statement

All participating IeDEA-SA sites obtained approvals from their local institutional review boards or ethics committees: Johannesburg/South Africa (ref. no. GP_202207_033), Lusaka/Zambia (ref. no. 2538-2022), Lilongwe/Malawi (ref. no. 22/02/2867), Chiure/Mozambique (ref. no. 89/CNBS/23), Masvingo/Zimbabwe (ref. no. MRCZ/A/2881). All study participants provided their written consent. In addition, the Cantonal Ethics Committee of Bern, Switzerland, approved the project (reference no. PB_2016-00273).

## Results

### Study participants

Among 1438 participants, 300 (21%) were enrolled in Malawi, 300 (21%) in Mozambique, 222 (15%) in South Africa, 361(25%) in Zambia and 255 (18%) in Zimbabwe. Table 1 shows their characteristics overall and by country: 363 (25%) of participants were ≤ 30 years, 963 (67%) males, 563 (40%) with HIV, 251 (17%) with a history of TB, 1214 (84%) with bacteriologically confirmed TB, and 59 (4%) with MDR-TB.

**Table 1:**
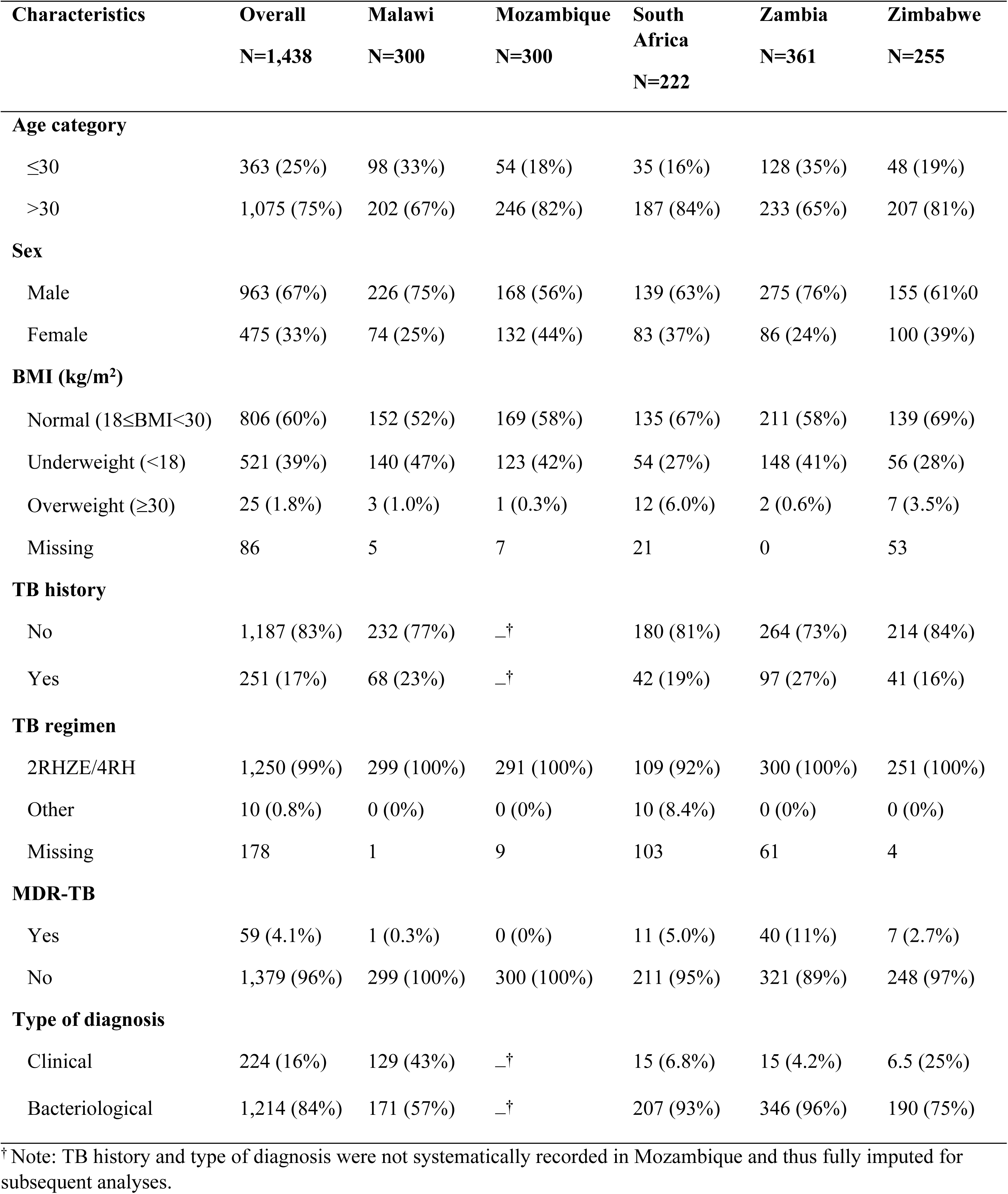
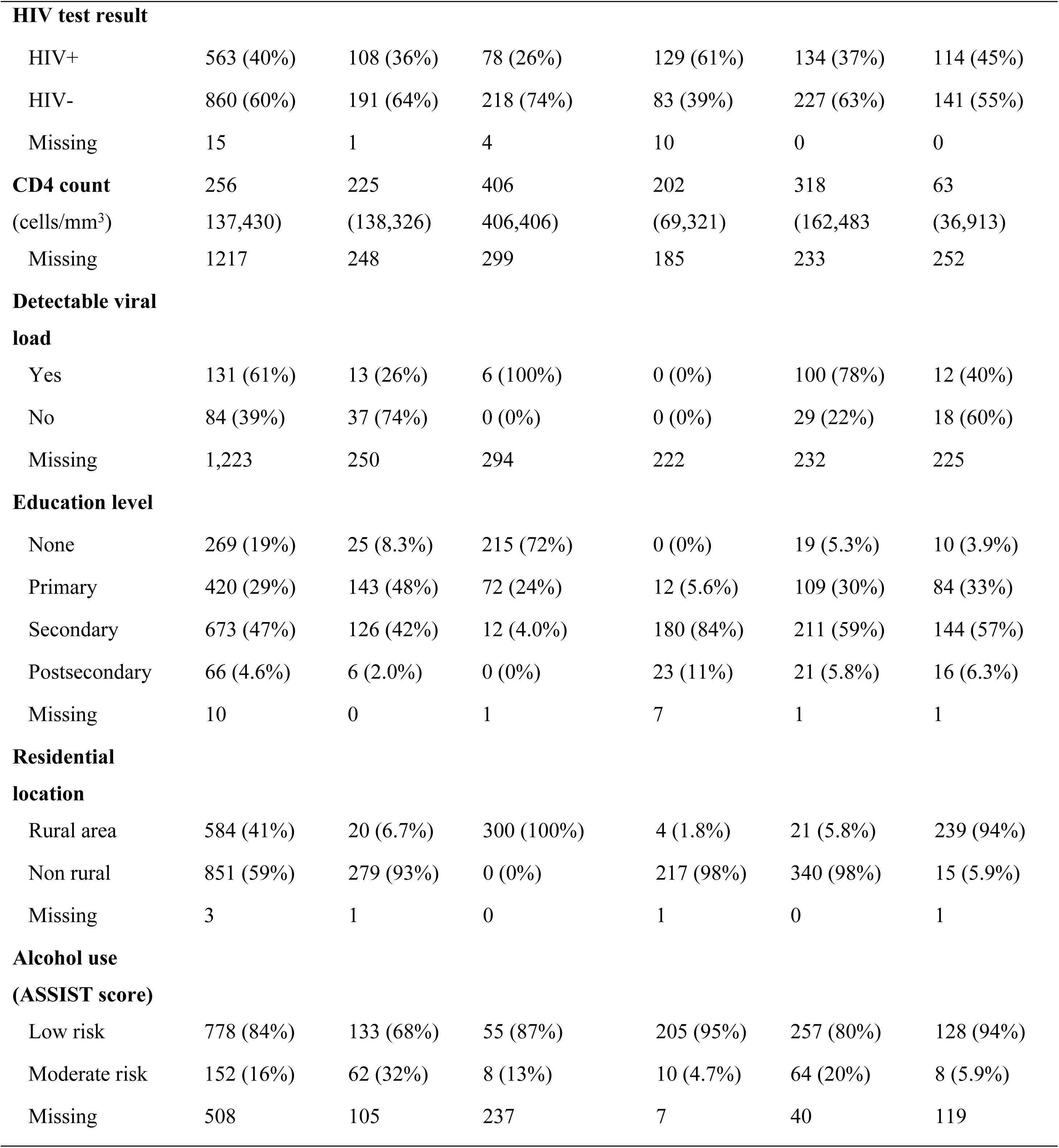
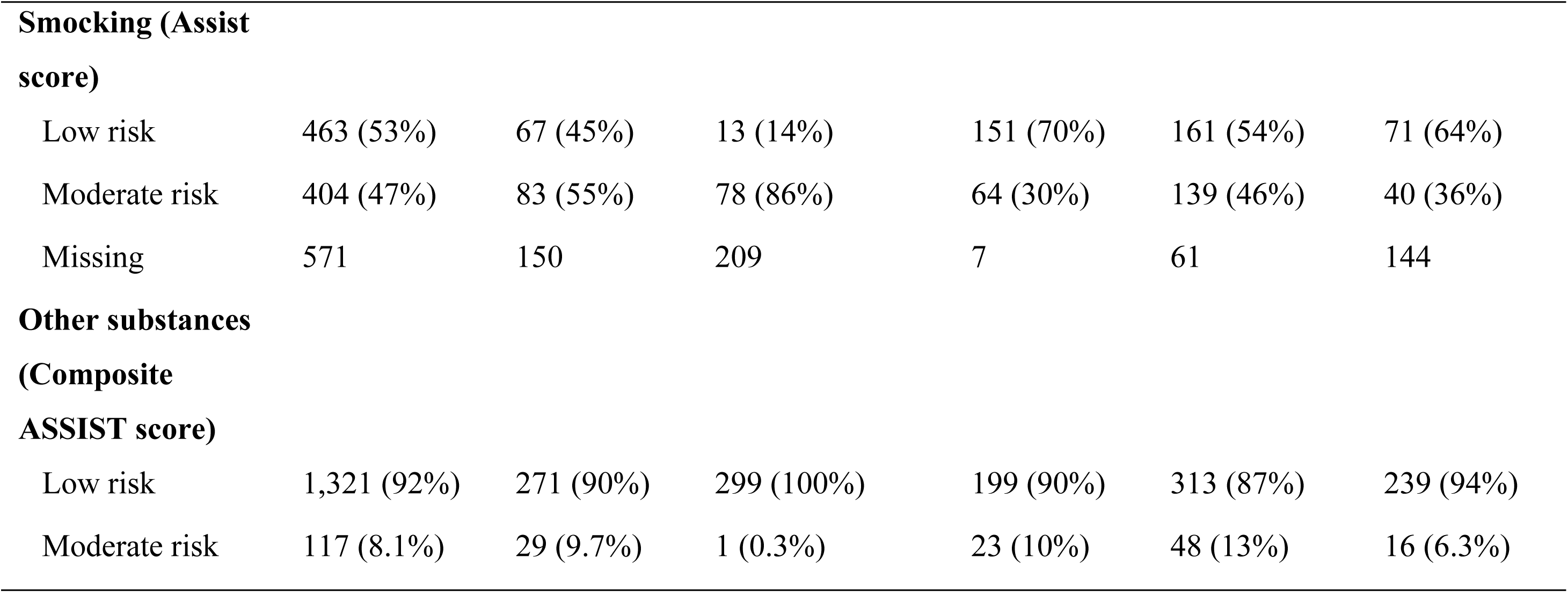
Patient and clinical characteristics of study participants overall and by country.

**Table 2:** TB treatment outcomes overall and by country.

| **Outcome n (%)** | **Overall** | **Malawi** | **Mozambique** | **South Africa** | **Zambia** | **Zimbabwe** |
| --- | --- | --- | --- | --- | --- | --- |
|  | N=1,438 | N=300 | N=300 | N=222 | N=361 | N=255 |
| **Treatment success** | 1,151 (80) | 278 (93) | 264 (88) | 138 (62) | 275 (76) | 196 (77) |
| Cured | 606 (53%) | 128 (46%) | 107 (41%) | 59 (43%) | 226 (82%) | 86 (44%) |
| Treatment completed | 545 (47%) | 150 (54%) | 157 (59%) | 79 (57%) | 49 (18%) | 110 (56%) |
| **Treatment unsuccessful** | 221 (15) | 21 (7) | 18 (6) | 67 (30) | 62 (17) | 53 (21) |
| Treatment failure | 3 (1.4) | 3 (14) | 0 (0%) | 0 (0%) | 0 (0%) | 0 (0) |
| Died | 89 (40) | 16 (76) | 11 (61) | 5 (7.5) | 22 (35) | 35 (66) |
| Loss to follow-up | 129 (58) | 2 (9.5) | 7 (39) | 62 (93) | 40 (65) | 18 (34) |
| **Other outcomes <sup>1</sup>** | 66 (5) | 1 (0) | 18 (6) | 17 (8) | 24 (7) | 6 (2) |
| Unknown | 49 (74) | 1 (100) | 18 (100) | 13 (76) | 14 (58) | 3 (50) |
| Transferred | 17 (26) | 0 (0) | 0 (0) | 4 (24) | 10 (42) | 3 (50) |

### TB treatment outcomes

The distribution of TB treatment outcomes is shown in Figure 1: 1151 (80%) successful, 89 (6%) deaths, 129 (9%) loss to follow-ups, 3 (<1%) treatments failures, 17 (1%) transfer-outs, and 49 (3%) unknown outcomes. The proportion of successful outcomes varied between countries (Table 1), with the highest proportion in Malawi (93%) and the lowest in South Africa (62%). Treatment success was lower for MDR-TB than non-MDR patients (56% versus 81%; Supplementary Figure 1). There was no significant difference in survival between participants with and without HIV (Supplementary Figure 2). Non-fatal serious adverse events were observed in 37 (3%) patients.

**Figure 1:**
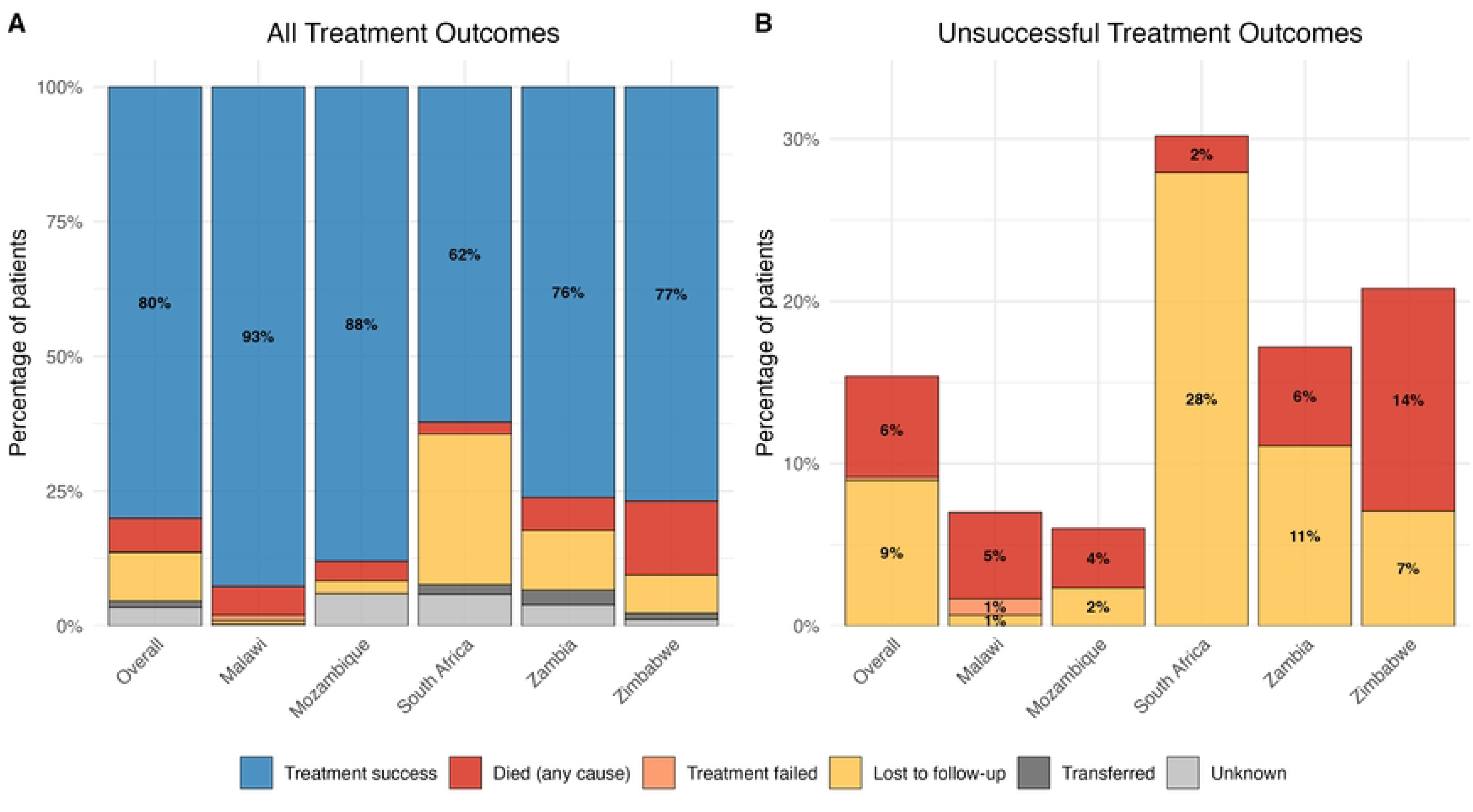
Distribution of (A) all and (B) unsuccessful TB treatment outcomes overall and by country.

### Predictors of TB treatment outcomes

Unsuccessful outcomes were more probable among patients with MDR-TB and among participants without formal education (Figure 2 and Supplementary Figure S3). Figure 3 shows the association of death and loss to follow-up with patient characteristics. The odds of death were lower for bacteriologically confirmed patients (aOR 0.45 95%-CI 0.25–0.80), patients with a BMI≥18 kg/m² (aOR 0.6, 95%-CI 0.36–0.99), and for participants with a secondary or higher level of education (aOR 0.3, 95%-CI 0.13–0.69). Moderate risk of smoking was associated with reduced odds of death (aOR 0.4, 95%-CI 0.13–0.92). MDR-TB was associated with a more than twofold increase in loss to follow-up (aOR 2.4, 95%-CI 1.17–4.97). Participants with secondary or higher and primary education were less likely loss to follow-up than participants without formal education (aOR 0.3, 95%-CI 0.14–0.89 and 0.3, 95%-CI 0.11–0.67, respectively). Non-fatal serious adverse events during treatment were more frequent among participants with HIV than participants without HIV (Supplementary Figure 4).

**Figure 2:**
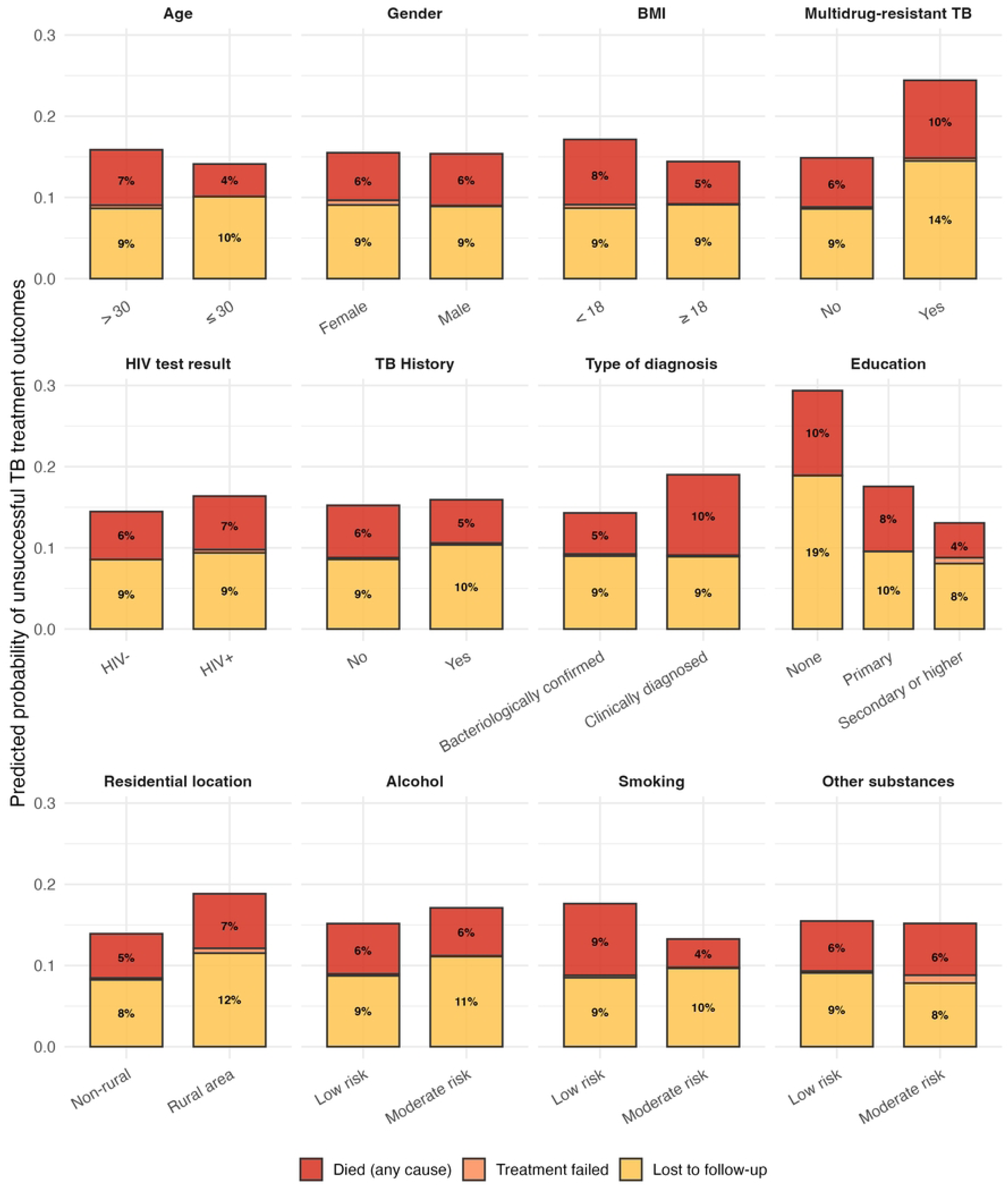
Probabilities of unsuccessful treatment outcomes by patient characteristics. Predicted probabilities were obtained by setting each covariate to a given level for the entire study population and averaging the resulting model-based predictions.

**Figure 3:**
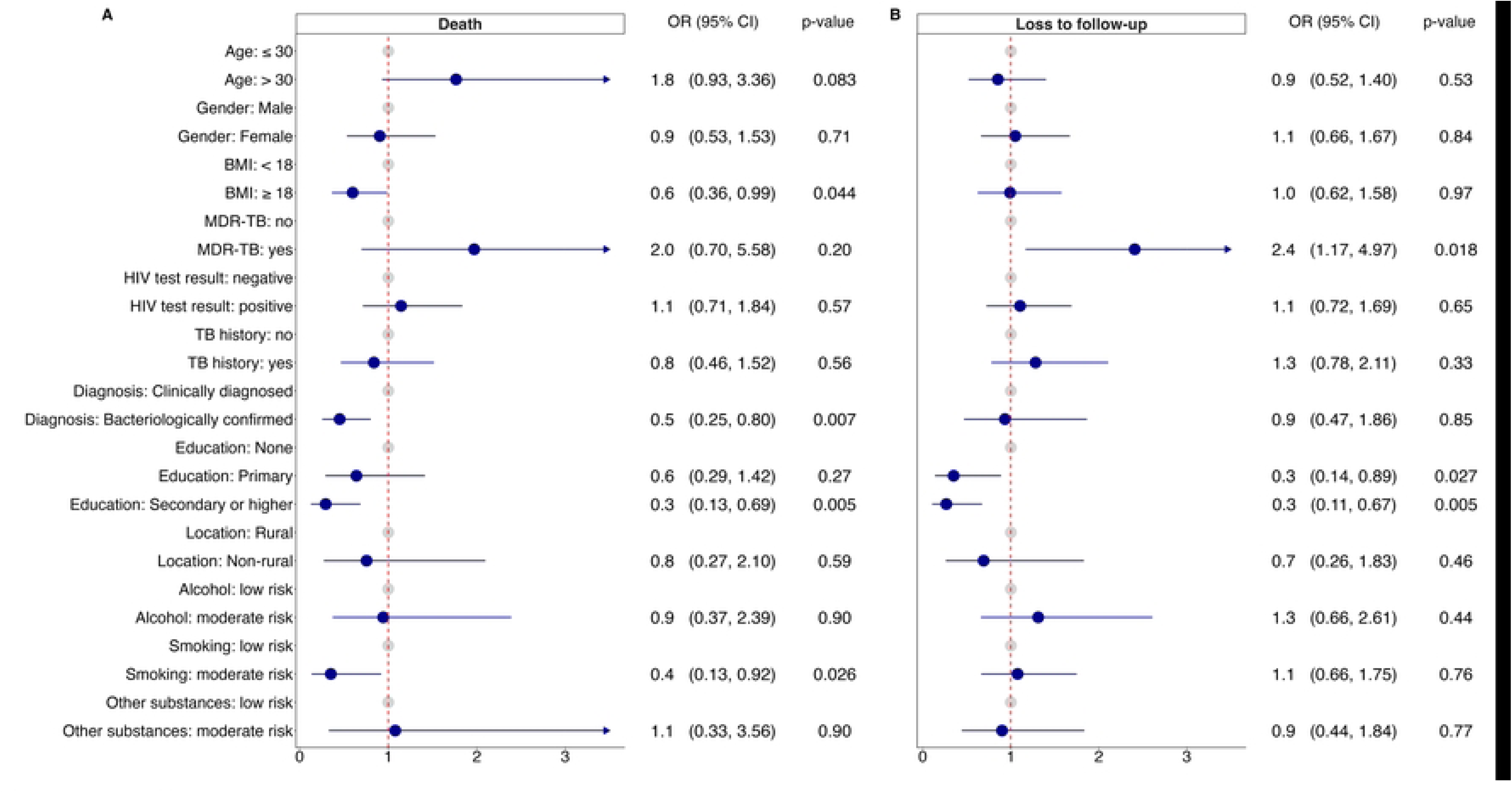
Patient factors associated with (A) death and (B) loss to follow-up. For death, unknown outcomes and loss to follow-ups were removed but transfer-outs were considered as “no deaths”. For loss to follow-up, unknown outcomes and deaths were removed but transfer outs were considered as “no loss to follow-ups”.

## Discussion

This study evaluated tuberculosis (TB) treatment outcomes and predictors of poor outcomes among patients treated for TB in Malawi, Zambia, Mozambique, Zimbabwe and South Africa. We enrolled 1438 participants of which 563 (40%) were HIV positive. We found that 80% of the participants had successful outcomes, 15% had unsuccessful outcomes (death and loss to follow-up) and 5% had unknown outcomes or were transferred out. MDR-TB was associated with more unsuccessful and higher education levels with more successful outcomes. Bacteriologically confirmed TB was associated with a reduced risk of death. Non-fatal serious adverse events during treatment were only rarely reported but more often among people with HIV.

The tuberculosis treatment success rate of 80% in our study cohort was below the WHO recommended TB target of ≥85% and below the global target for the end TB strategy of ≥90% by 2035 (12). However, it is close to the 79% rate found across Africa in a systematic review and metanalysis (6). Compared to region-specific estimates, the success rate in our cohort was lower than in Galkayo Puntland, Somalia (85%), East Wollega, Western Ethiopia (92%), Bosaso TB hospital, Somalia (89%), Zomba, Malawi (89%) (13–16), but higher than in Kyangwali Refugee Settlement, Uganda (55%) (17). Differences between studies might be due to differences in socioeconomic status, education level, access to health facilities, geographic location, study design, duration and sample size. There might also be an overall decline in TB treatment success due to disruptions in TB care during the COVID-19 pandemic, specifically as a result of reductions in national funding or development aid for TB treatment programs, which affected TB prevention and control activities, timing for TB diagnosis and treatment, care seeking behaviors and healthcare utilization even after the COVID 19 pandemic (18).

We found that about 15% of the participants experienced unsuccessful treatment outcomes, which consisted of 6% deaths, 9% loss to follow-ups and <1% treatment failures. This contrasts with 21% unsuccessful outcomes in Africa found by the systematic review and metanalysis, consisting of 10% deaths, 10% loss to follow-ups and 1% to treatment failures (6). A study conducted at Mildmay hospital in Uganda had a 10.2% mortality estimate in 2022 which was higher than our findings (19). Our rate for unsuccessful outcomes was within the range set by the WHO in 2020 of 15% among new TB cases and 24% among patients coinfected with HIV (12), although it was higher than the 10% end TB strategy target. Providing patient education and counselling, strengthening direct observed therapy, patient tracing, setting up reminders of TB drug collection dates, and drug intake times could improve TB treatment outcomes (20,21).

Bacteriologically confirmed TB diagnosis was associated with a 50% lower risk of death. This is in line with a systematic review which found that people with clinically diagnosed TB were 50% more likely to die, possibly because other important medical conditions were also not diagnosed or due to the delay of optimal treatment (22). The proportion of clinically diagnosed TB was 40% in the systematic review, compared with only 20% in our study. Conversely, the proportion of bacteriologically confirmed patients in our study was higher than in the WHO global TB report (80% versus 62%) (23). The lower proportion of clinically diagnosed TB patients in our cohorts could thus have positively impacted the TB success rate. Note that higher proportions of clinically diagnosed TB were seen in patients with HIV, hospitalized patients and among extrapulmonary TB patients (22).

Study participants with a secondary or higher education had a lower risk of death than those without a formal education. This is consistent with a study done in Columbia that showed higher TB mortality among participants with the lowest education level (24). However, a study that was done in Durban, South Africa found no association between education and TB treatment outcomes among participants with HIV (25). Differences in study designs, study settings and sample size might have contributed to variations. Level of education is linked to health literacy and income seeking behavior. Health literacy is important for tuberculosis prevention through improved knowledge, early TB detection through health seeking behaviors, and greater adherence to the course of treatment leading to improved TB treatment outcomes (28). Educated participants are also more likely to have a higher socioeconomic status, better living conditions and improved nutrition, which may altogether contribute to more successful TB treatment outcomes (6).

Moderate tobacco smoking was surprisingly associated with lower risk of death. Another study that was conducted in West Africa showed no significant association between smoking and unsuccessful TB treatment outcomes (26). However, there are many studies that have reported an association between smoking and mortality. A systematic review and meta-analysis that was conducted in Africa showed that smoking increased likelihood of poor TB outcomes by 30-75% (27). Other systematic reviews and meta-analysis studies have also shown association of smocking with TB treatment outcomes (28,29). We speculate that increased awareness of health risks as well as earlier and more frequent hospital visits associated with smoking-related medical conditions could have led to improved TB diagnosis and care which mediated the effect on TB treatment outcomes in this study.

MDR-TB had more than doubled the risk of death and loss to follow-up. MDR-TB had a 56% probability of treatment success, which is similar to the 50% found in a study in Ethiopia (30). These estimates are before newer oral regimens became available, so that the longer duration and toxicity of older MDR-TB drug regimens may have been linked to increased loss to follow-up (31). Furthermore, a study that was conducted in Surabaya, Indonesia, found that negative attitudes towards treatment, lack of social support, low socioeconomic status and lack of satisfaction with health services were associated with loss to follow-up among MDR patients (32). Other factors that have shown association to poor TB treatment outcomes among MDR patients were comorbidities such as HIV and malnutrition as well as delayed TB diagnosis and treatment initiation (30,33–35).

Our study is subject to several limitations. First, the participating IeDEA sites are not fully representative of the national health infrastructure for tuberculosis and HIV care in each country, potentially limiting the external validity of our findings. However, the use of a multi-centric design spanning diverse geographic locations ensures that the data broadly reflect the clinical realities of high-burden settings in Sub-Saharan Africa. Second, we acknowledge that observed variations between country cohorts may not exclusively reflect epidemiological differences but may also stem from operational heterogeneity in study implementation and data abstraction by local site investigators. To account for this unmeasured site-level variance, we incorporated country-specific random and fixed effects into our multivariable association analyses. Third, longitudinal attrition remains a challenge, particularly given the high rates of circular migration and population mobility characteristic of the Southern African region. Fourth, while loss to follow-up was categorized as a distinct treatment outcome, the true clinical endpoints remain unknown. Finally, data completeness varied across covariates; to mitigate bias associated with missingness, these covariates were imputed and estimation results were pooled across multiple imputed datasets.

In conclusion, we found that successful treatment outcomes were below WHO targets. Loss to follow-up and death remained high despite integration of TB care into HIV programs. MDR-TB was associated with higher loss to follow-up, while secondary/higher formal education reduced loss to follow-up and risk of death. Microbiologically diagnosed TB was also associated with a reduced risk of death. Altogether, this suggests that having a definitive TB diagnosis through microbiological confirmation and instituting retention strategies is critical for improving TB treatment outcomes. Future TB research should focus on definitive TB diagnosis, development of shorter, safer, and effective TB regimes and strategies of retaining MDR-TB patients in care.

## Acknowledgements

We would like to thank all the participating clinics and all study participants. We are also grateful to the local study teams for their diligent daily work.

**Centre for Infectious Disease Research in Zambia, Lusaka, Zambia:**

Esau Banda, Carolyne Bolton, Carol Chileshe Chishimba, Mable Chungulo, Jackson Daka, Mary Kanyanta, Paxina Katunga, Guy Kayeye Muula, Mary Mwale, Ethel Muyanga, Alice Miyanda, Fiona Mureithi, Sylvia Musonda, Judith Mwanza, Suwilanji Nalungwe, Cynthia Nayame, Veronica Phiri, Kenan Simumba, Vivian Tonga.

**Themba Lethu Clinic, Johannesburg, South Africa:**

Tokologo Chiloane, Denise Evans, Nosihle Thembelihle Fairhope Hlophe, Funeka Jara, Nelly Jinga, Sinethemba Madlala, Cassandra Mbanje, Thuso Enock Malatsi, Betty Matome, Violet Molepo, Nkamoheleng Mokhesi, Nompumelelo Mthupa, Melda Musina, Nkateko Lebogang Ngolele, Pertunia Rakgoho, Clive Ramushu, Nomaphelo Sokatsha, Hazel Tau.

**Lighthouse Clinic, Lilongwe, Malawi:**

Mervis Bonongwe, Joseph M. Chintedza, Shadreck Chipapi, Geldert Chiwaya, Jane Chiwoko, Layout Gabriel, Jessie Hau, Jacqueline Huwa, Mike Kalulu, Aubrey Kudzala, Angella Mtambalika, Erick Mtemang’ombe, Blessings Mwandira, Bernard Mzikalenga, Peter Muula, Richard Nyambalo, Mirriam Ndhlovu Soko, Ethel Rambiki, Agness Thawani.

**SolidarMed Mozambique:**

Mussa Aly, Amina Fernando Ansuma, Arminda Bernardo Arabe, Saide Borges, Cardoso Estevao, Belermino Eugenio, Edson Fernandes, Alfredo Joao, Anastacia Lidimba, Murane Olaide Malimo, Belinha Jose Molde, Karolin Pfeiffer, Magido Sabune, Thomas Vandamme.

**SolidarMed Zimbabwe:**

Joseph Bishi, Brian Dhlandara, Mutsa Gwatinyanya, Cuthbert Magande, Lovemore Masukume, Shamaine Mhondiwa, Sarudzai Muzorori, Thalia Mungwari, James Nyamutsamba, Karolin Pfeiffer, Kumbirai Pise Quarter, Laura Ruckstuhl, Tandiwe Sithole, Amadeus Shamu, Tauro, Takayidza.

## Authors’ contributions

Conception and design: MN, LW, JH, AT, NB, and LF. Data collection: MN, GC, JC, AK, JH, AT, GM, DE, IR, CK, FM, NJ, AM, MB, and GG. Data analysis: NB and LW. Wrote the first draft: MN, LW, JH, AT, GC, NB, and LF.

## Data availability statement

Complete data for this study cannot be posted in a supplemental file or a public repository because of legal and ethical restrictions. The Principles of Collaboration of this multi-national consortium and the regulatory requirements of the different countries’ IRBs require the submission and approval of individual project concept sheets that describe the planned analyses. Specifically, while the data held by the IeDEA-SA consortium may be available to other investigators, the proposed use must be based on a concept note that is approved by the regional Steering Groups (contact: https://www.iedea-sa.org/contact-us/).

---

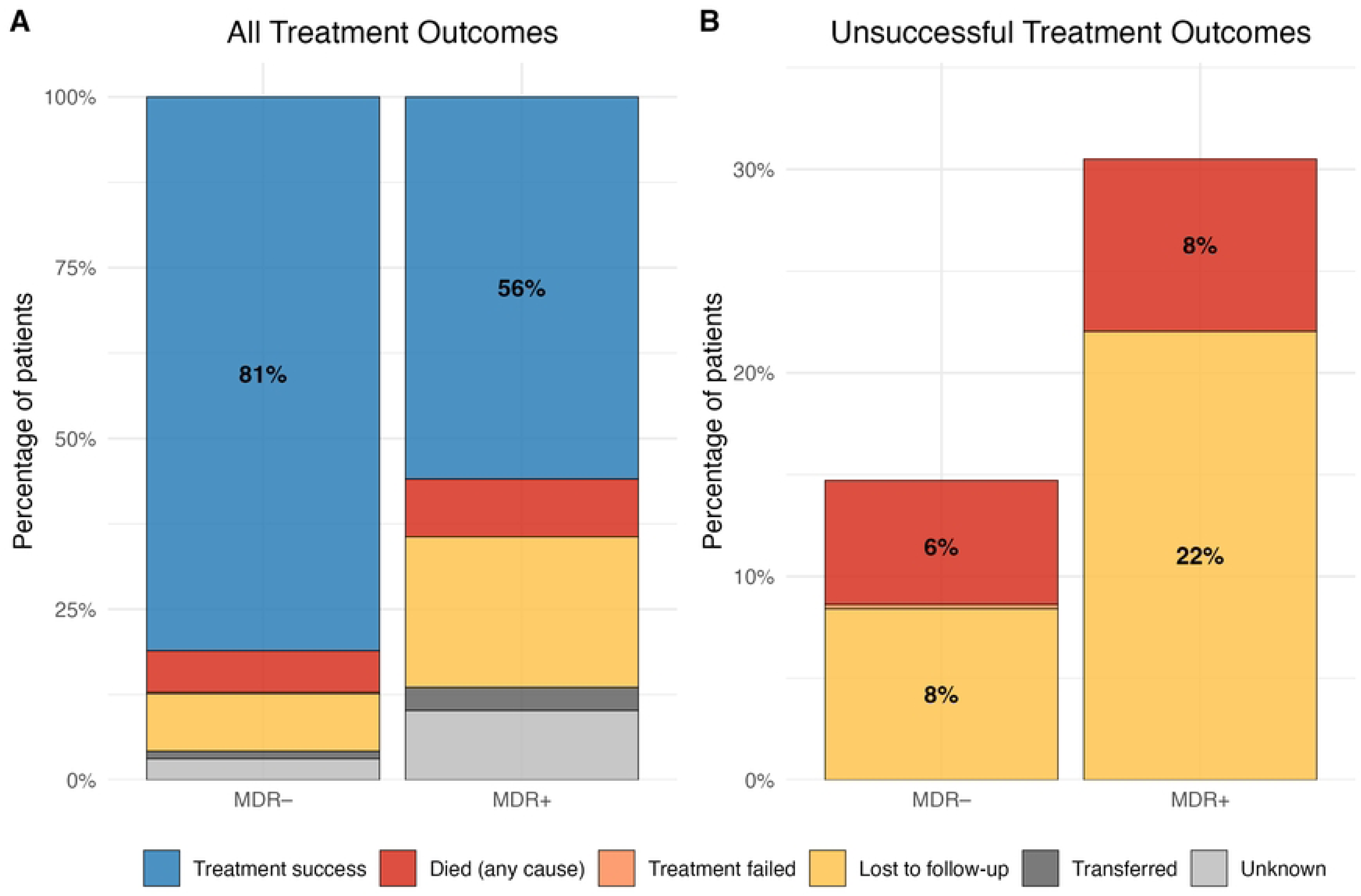

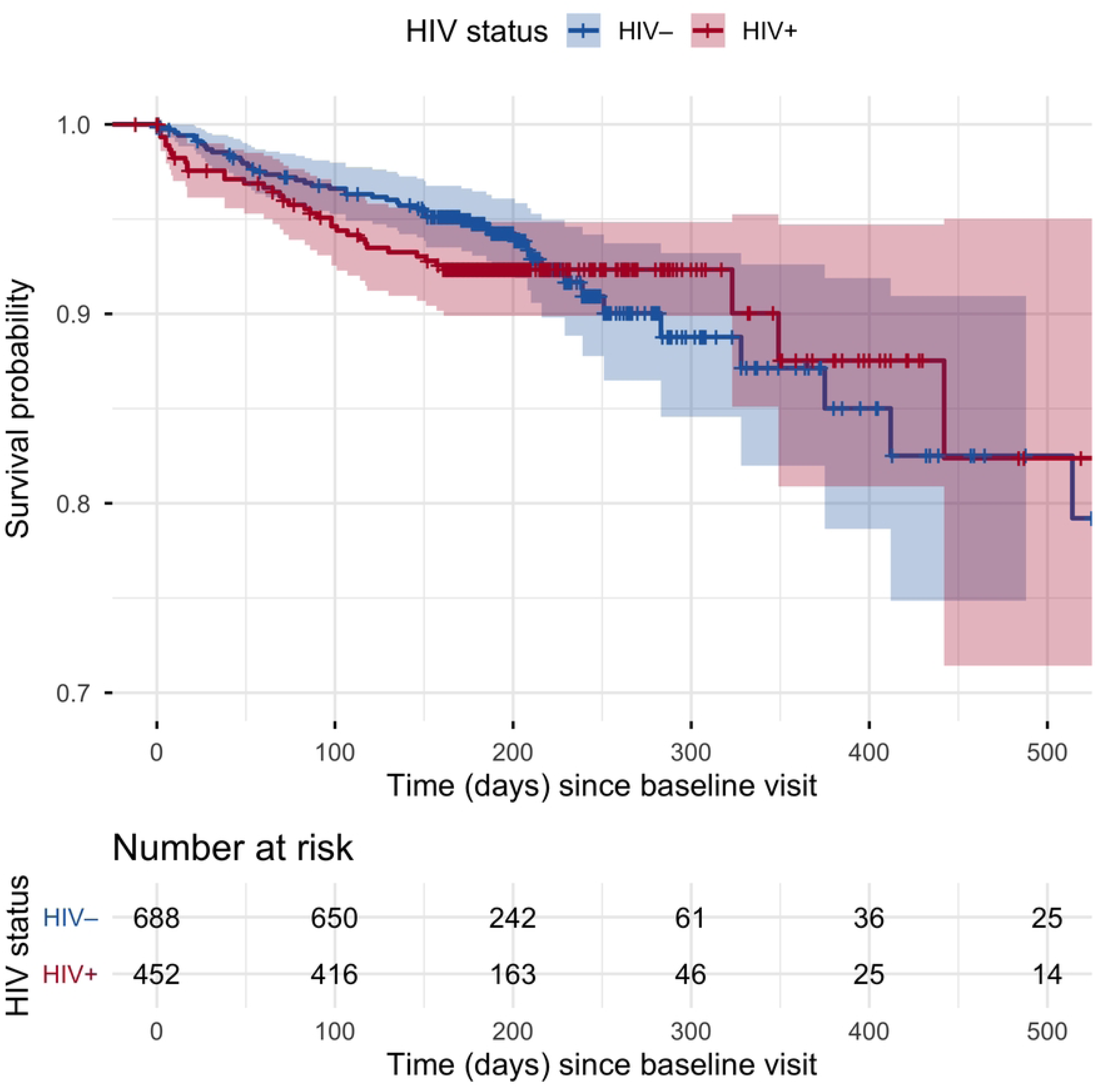

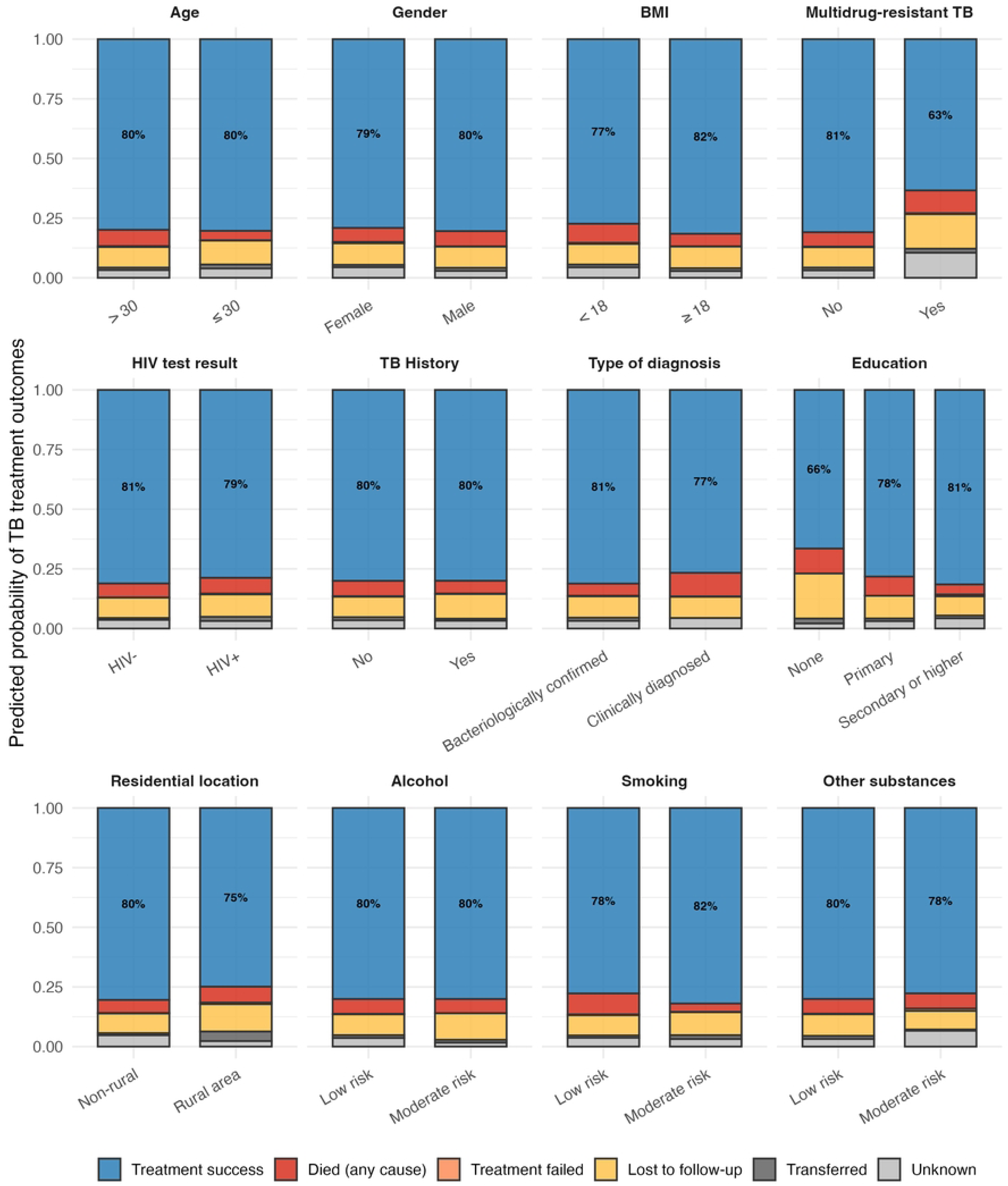

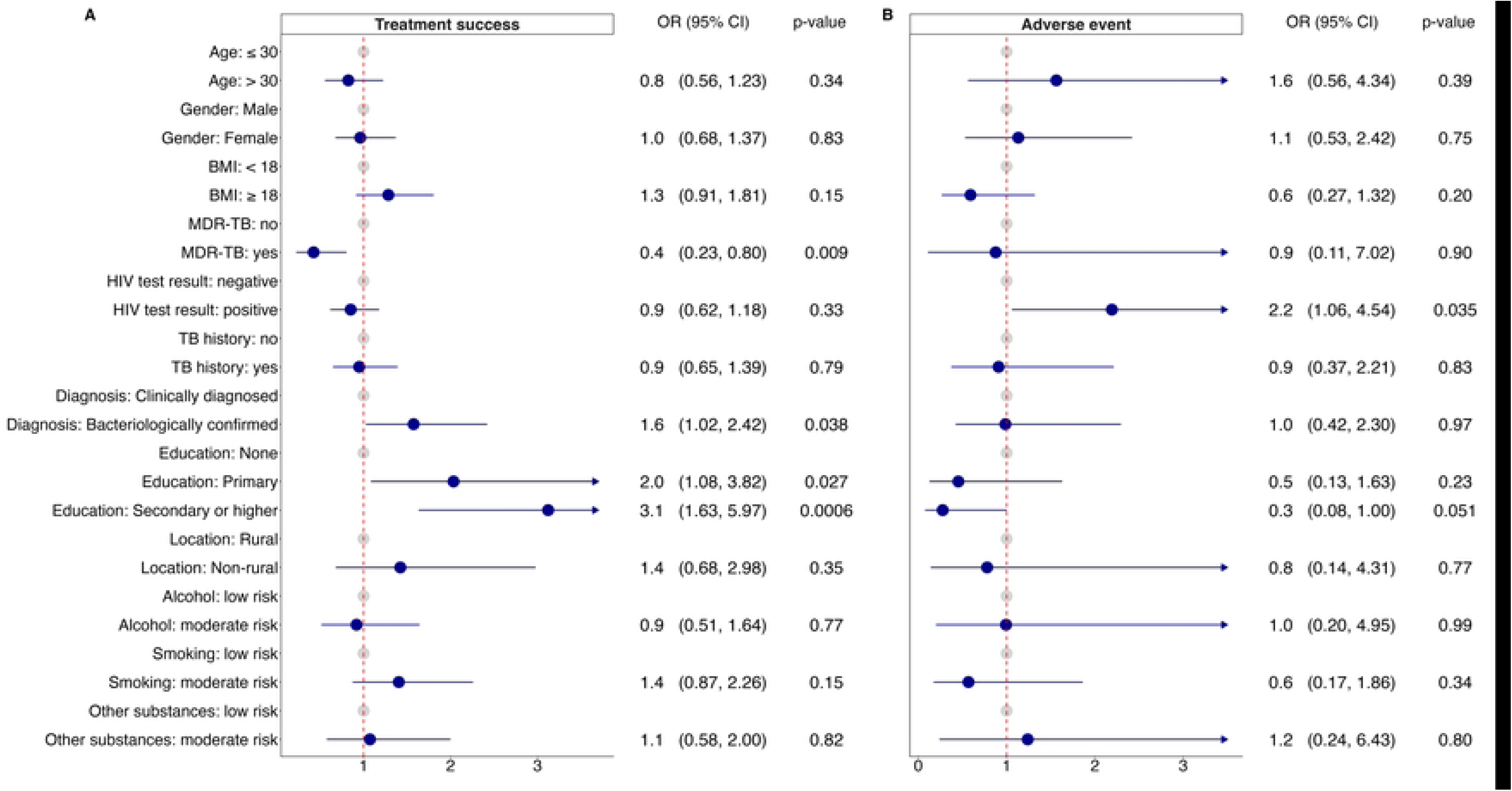

